# Sarilumab use in severe SARS-CoV-2 pneumonia

**DOI:** 10.1101/2020.05.14.20094144

**Authors:** Elisa Gremese, Antonella Cingolani, Silvia Laura Bosello, Stefano Alivernini, Barbara Tolusso, Simone Perniola, Francesco Landi, Maurizio Pompili, Rita Murri, Angelo Santoliquido, Matteo Garcovich, Michela Sali, Gennaro De Pascale, Maurizio Gabrielli, Federico Biscetti, Massimo Montalto, Alberto Tosoni, Giovanni Gambassi, Gian Ludovico Rapaccini, Amerigo Iaconelli, Lorenzo Zileri Del Verme, Luca Petricca, Anna Laura Fedele, Marco Maria Lizzio, Enrica Tamburrini, Gerlando Natalello, Laura Gigante, Dario Bruno, Lucrezia Verardi, Eleonora Taddei, Angelo Calabrese, Francesco Lombardi, Roberto Bernabei, Roberto Cauda, Francesco Franceschi, Raffaele Landolfi, Luca Richeldi, Maurizio Sanguinetti, Massimo Fantoni, Massimo Antonelli, Antonio Gasbarrini, on behalf of the GEMELLI AGAINST COVID^

## Abstract

**Importance:** Interleukin-6 signal blockade has shown preliminary beneficial effects in treating aberrant host inflammatory response against SARS-CoV-2 leading to severe respiratory distress.

**Objective:** to describe the effect of off-label intravenous use of Sarilumab in patients with severe SARS-CoV-2-related pneumonia.

**Design:** Observational clinical cohort study.

**Setting:** Fondazione Policlinico Universitario A. Gemelli IRCCS as Italian Covid reference center.

**Participants:** Patients with laboratory-confirmed SARS-CoV-2 infection and respiratory distress with PaO_2_/FiO_2_ ratio<300 treated with Sarilumab between March 23^rd^ – April 4th, 2020. Date of final follow-up was April 18, 2020.

**Main outcomes and measures:** We describe the clinical outcomes of 53 patients with SARS-CoV-2 severe pneumonia treated with intravenous Sarilumab in terms of pulmonary function improvement or Intensive Care Unit (ICU) admission rate in medical wards setting and of live discharge rate in ICU treated patients as well as in terms of safety. Each patient received Sarilumab 400 mg administered intravenously on day 1, with eventual additional infusion based on clinical judgement, and was followed for at least 14 days, unless previously discharged or dead. No glucocorticosteroids were used at baseline.

**Results:** Of the 53 SARS-CoV-2^pos^ patients receiving Sarilumab, 39 (73.6%) were treated in medical wards (66.7% with a single infusion) while 14 (26.4%) in ICU (92.6% with a second infusion). The median PaO_2_/FiO_2_ of patients in the Medical Ward was 146(IQR:120-212) while the median PaO_2_/FiO_2_ of patients in ICU was 112(IQR:100-141.5), respectively.

Within the medical wards, 7(17.9%) required ICU admission, 4 of whom were re-admitted to the ward within 5-8 days. At 19 days median follow-up, 89.7% of medical inpatients significantly improved (46.1% after 24 hours, 61.5% after 3 days), 70.6% were discharged from the hospital and 85.7% no longer needed oxygen therapy. Within patients receiving Sarilumab in ICU, 64.2% were discharged from ICU to the ward and 35.8% were still alive at the last follow-up. Overall mortality rate was 5.7% after Sarilumab administration: 1(2.5%) patient died in the Medical Ward whilst 2(14.2%) patients died in ICU, respectively.

**Conclusions and relevance:** IL-6R inhibition appears to be a potential treatment strategy for severe SARS-CoV-2 pneumonia and intravenous Sarilumab seems a promising treatment approach showing, in the short term, an important clinical benefit and good safety.

**Key points:** *Question:* SARS-CoV-2 infection remains a disease with many unknown aspects for which there are no therapies with proven efficacy. To date, it is recognized that COVID-19 disease may lead to the development of a cytokine storm for which drugs as IL-6R inhibitors may have beneficial effect.

*Findings:* In this observational clinical study, we reported the efficacy and safety of intravenous Sarilumab use in SARS-CoV-2 severe pneumonia with a global resolution rate of 83.0% (89.7% in medical wards and 64.3% in ICU) and an overall mortality rate of 5.7%.

*Meaning:* IL-6R-inhibition is an effective approach for severe SARS-CoV-2 pneumonia and intravenous Sarilumab is a promising treatment approach leading to an important clinical benefit and good safety in the short term.

## INTRODUCTION

On December 31, 2019, an outbreak of pneumonia of unknown etiology was reported in the Chinese city of Wuhan and 10 days later the Chinese Center Disease Control (CDC) identified the causative agent of this infection, a new coronavirus called SARS-CoV-2.^1^ On February 21, 2020, the first Italian patient with SARS-CoV-2 disease was admitted to the Codogno Hospital^2^ and over the next weeks there has been an exponential growth of cases in our country. To date, Italy is globally one of the most affected countries, with 205.463 confirmed cases, 27.967 deaths and 75.945 people declared recovered at April 30^th^ 2020.^3^

Although 81% of SARS-CoV-2^pos^ patients develop a mild disease, severe pneumonias requiring hospitalization are documented in about 14% of cases and the remaining 5% needs admission to Intensive Care Unit (ICU).^4-7^ Severe clinical scenario mimics a real “cytokines storm” similar to that occurring in Car-T leukemia treatment^8-9^ In this context, Interleukin-6 (IL-6) is considered a key cytokine mediating the aberrant activation of immune cells arosing as a putative mediator of SARS-CoV-2 induced cytokine storm.^10^ After FDA approval, anti-IL6R (i.e. Tocilizumab) has been adopted to treat SARS-CoV-2 severe pneumonia (www.clinicaltrials.gov). Due to its increased use and limited supply, Tocilizumab became rapidly unavaiable in Italy and Sarilumab, an alternative IL-6R inhibitor approved for Rheumatoid Arthritis (RA),^11^ has been chosen by the Covid-19 Gemelli Task Force to treat patients with severe SARS-CoV-2 pneumonia.

In the present study, we describe the effect of the off-label intravenous use of Sarilumab in patients with severe SARS-CoV-2-related pneumonia. In particular, the study endpoints were to assess (i) the impact of Sarilumab for the treatment of severe SARS-CoV-2 pneumonia in terms of pulmonary function improvement and prevention of ICU admission in a ward setting as of live discharge rate in patients treated in ICU care; (ii) safety and (iii) putative biological and clinical parameters related to improvement or ICU transfer after Sarilumab.

## METHOD

### Patients enrollment

From March 14^th^ 2020, Fondazione Policlinico Universitario A. Gemelli-IRCCS became a referral hospital (COVID-2 Hospital) for SARS-CoV-2 infected people. Hospitalized patients with severe SARS-CoV-2 pneumonia, defined as SARS-CoV2 infection confirmed by RT-PCR assay, interstitial pneumonia at imaging (chest X-Ray or CT scan), impairment of respiratory function (PaO_2_/FiO_2_ ratio<300), rapid worsening of the respiratory condition or need for ICU admission were considered eligible to anti-IL6R therapy. The shortage of Tocilizumab together with the increasingly number of patients with progressive respiratory symptoms led the multidisciplinary team made of Immuno-rheumatologists, Infectivologists, Pneumologists and Covid-wards clinicians to use Sarilumab, according to a shared clinical-pharmacological protocol (Prot.n.926 approved by the Ethics Comittee of the Fondazione Policlinico Universitario A. Gemelli-IRCCS, Università Cattolica del Sacro Cuore). Patients enrollment started on March 23^rd^ till April 4^th^, 2020, and data collection continued until April 18^th^, 2020, to ensure at least 14 days follow-up for each patient. Sarilumab treatment was scheduled as follows: 400 mg intravenously on day 1, according to the published Italian Medicines Agency (AIFA) protocol (final injectable solution was obtained combining 2 Sarilumab 200 mg prefilled syringes mixed in 100 ml 0.9% sodium chloride solution for intravenous use). In case of clinical worsening or unchanged status, a repeated dose of Sarilumab 400 mg i.v. was administered. Each patient or next of kin provided informed consent. Concomitant therapies were set for all patients at the time of confirmed positive naso-pharyngeal swab for SARS-CoV-2 upon entering the Hospital, unless differently required by clinical situation, as follow: lopinavir/ritonavir 400/100 mg BID or darunavir/ritonavir 800/100 mg QD, orally); Hydroxychloroquine (400 mg BID for the first day, followed by 200 mg BID); Azithromycin combined at the dose of 500 mg orally or intravenously. Moreover, prophylactic dose heparin subcutaneously (4000-6000 UI QD) was added. In case of ICU admission, the Intensivist may use GC as suggested by the Society of Critical Care Medicine.^12^

### Clinical, inflammatory and immunological parameters collection

For each patient, demographic, clinical and immunological data were collected at study entry and after 24h, 3, 5, 7 and 14 days of follow-up and clinical outcome was reported at last follow-up (ongoing hospitalization, hospital discharge or death, respectively). In particular, peripheral arterial blood oxygen status was checked and respiratory status (PaO_2_/FiO_2_ and SpO_2_ at rest) together with oxygen-support requirement (room air, Ventimask, CPAP, noninvasive ventilation-NIV, high flow nasal cannula-HFNC), laboratory data were recorded at each timepont and at last follow-up. At the same timepoints, peripheral blood was collected for the assessment of IL6 plasma levels using ELISA assay (Multi-cytokine test for ELLA-Bio-Techne, Minneapolis).

### Clinical improvement and outcomes assessment

Clinical improvement was defined as the reduction of oxygen-support requirement or improvement of the PaO_2_/FiO_2_ ratio from baseline, as discharge rate and/or no longer need of oxygen supplementation. In addition, the need of ICU admission for patients treated in medical wards (and the subsequent discharge/exitus outcome) and the discharge towards medical wards for ICU-treated patients were evaluated.

### Statistical analysis

Statistical analysis was performed using SPSS V.20.0 (SPSS. Chicago, Illinois, USA) and Prism software (GrapAHad, San Diego, California, USA). Categorical and quantitative variables were described as frequencies, percentage and Median (25-75 interquartile range, IQR). Data on demographic and clinical parameters were compared between patients by the non-parametric Mann-Whitney U test or χ^2^ test, as appropriate. Univariate analysis was conducted to assess adequate event frequency between the outcomes and the candidate predictors. Predictors with a p<0.05 entered within the multivariable logistic regression analysis.

A Receiver Operating Characteristic (ROC) curve analysis of clinical and laboratory parameters differentially distributed among SARS-CoV-2^pos^ patients based on the considered outcome was performed to obtain relevant threshold for the prediction of “clinical improvement at 3 days after Sarilumab treatment” and of “transfer to ICU”, respectively. The optimal cut-off points were determined to yield the maximum corresponding sensitivity and specificity. Moreover, Kaplan Meyer analysis was performed to estimate the probability of “transfer to ICU” in SARS-CoV-2^pos^ patients based on the previously identified cut-off values of clinical and laboratory parameters.

Finally, a nomogram was built to distinguish patients with the outcome (transfer to ICU) and those without. The performance of the nomogram was assessed by discrimination and calibration. The discriminative ability of the model was determined by the area under the ROC curve, which ranged from 0.5 (no discrimination) to 1 (perfect discrimination). The model was developed and validated. The statistical analyses and graphics were performed using the *rms* statistical packages of R 3.5,3 (The R Foundation for Statistical Computing, Vienna, Austria). For all the analyses, a p<0.05 was considered statistically significant.

## RESULTS

### Baseline demographic, clinical and inflammatory characteristics of SARS-CoV-2^pos^ patients treated with Sarilumab

Fifty-three patients with severe SARS-CoV-2-related pneumonia were enrolled, of whom 47(88.7%) were male with an age range of 40-95 years (median 66 years), 33(62.3%) were overweight/obese and 34(64.2%) had at least one comorbidity (**Supplementary Tables 1-2**).

Considering concomitant therapies, 37(69.8%) were treated with darunavir/ritonavir, 13(24.5%) with lopinavir/ritonavir while 3(5.7%) received no antiviral treatment. Moreover, 50(94.3%) were treated with hydroxychloroquine, 45(74.9%) were under prophylactic dose heparin and 29(54.7%) received azythromicin. No patients were taking GC at baseline. Among the whole cohort, 39(73.6%) patients were treated in medical wards while 14(26.4%) received their first Sarilumab dose in ICU or within the 24h of ICU admission (**Supplementary Tables 1-3**).

**Figure 1A and 2A** show individual clinical course of SARS-CoV-2^pos^ patients in medical wards and ICU, respectively. Twenty-six (66.7%) patients belonging to medical wards received a single course of Sarilumab, while 13(33.3%) received a second dose at a median interval of 3 days (range 1-11 days), based on the clinical course (**Figure 1A**). Conversely, 13(92.9%) ICU-treated patients received two Sarilumab infusions [median interval: 2.5 days (range 1-5 days)] (**Figure 2A**). The dose of Sarilumab was reduced in two patients: one HIV^pos^ and HBV^pos^ patient under chronic specific treatment received two courses of Sarilumab (200 mg each, 3 days apart), experiencing clinical improvement at day 3 and discharged from the hospital at day 14; another 95 years-old patient received a single infusion of Sarilumab 200 mg, showing PaO_2_/FiO_2_ ratio improvement from 134 to 270 within 24h and oxygen support reduction from 50% to 24% in 7 days. Two patients received a reduced dose of Sarilumab at re-treatment, due to mild neutropenia development after the first dose.

**Figure 1A-E.**
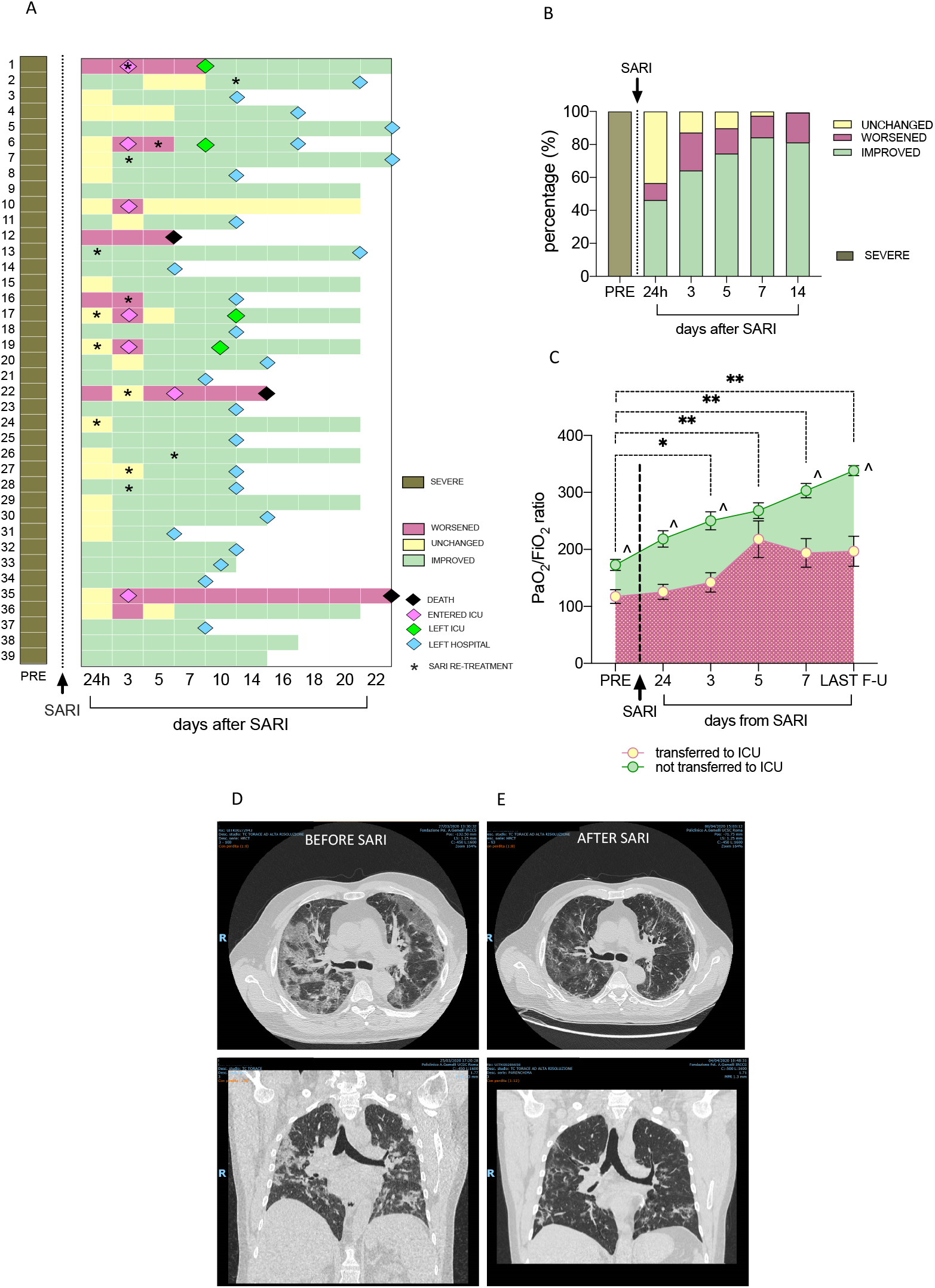
Clinical course of SARS-CoV-2^pos^ patients treated with Sarilumab in medical wards. **A)** Individual disease course of SARS-CoV-2^pos^ patients in medical wards. Each individual disease course is depicted in a line from pre-treatment to maximum 22 days after Sariumab administration; Pink diamonds refer to transfer towards ICU; Green diamonds refer to transfer back from ICU to medical wards; Light blue diamonds refer to discharge from the hospital and black diamond refers to death. Black stars indicate repetition of infusion of Sarilumab during the follow-up. **B)** Rate of clinical response to Sarilumab in SARS-CoV-2^pos^ patients within 14 days follow-up (24h, 3,5,7 and 14 days) in medical wards. **C)** PaO_2_/FiO_2_ ratio in SARS-CoV-2^pos^ patients treated with Sarilumab based on their need to be transferred to ICU during the follow-up; comparison between baseline vs 3 days *p<0.001; **p<0.0001 refer to comparison between baseline vs 5, 7 days and last follow-up respectively; ^^^p<0.01 for each time-point comparing patients trasferred to ICU vs patients not transferred to ICU after Sarilumab treatment. **D-E)** Example photos of pulmonary CT-scan of SARS-Cov-2^pos^ patients before (D) and after (E) Sarilumab treatment.

**Figure 2A-B.**
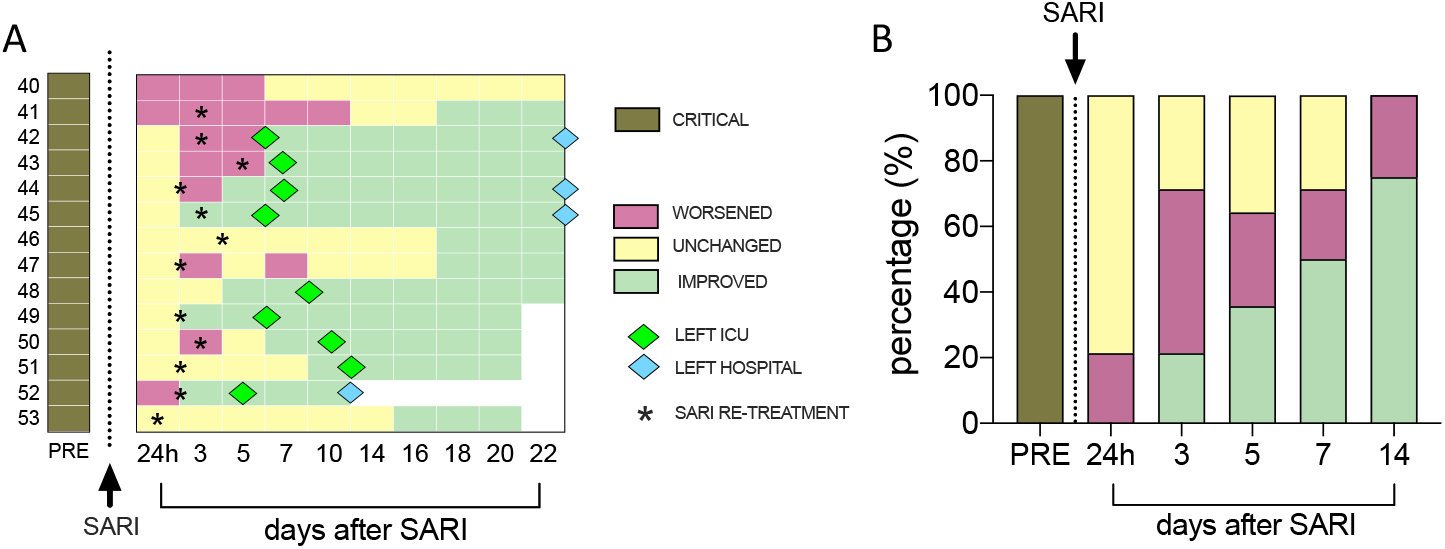
Clinical course of SARS-CoV-2^pos^ patients treated with Sarilumab in the ICU setting. A) Individual disease course of SARS-CoV-2^pos^ patients in ICU. Each individual disease course is depicted in a line from pre-treatment to maximum 22 days after Sarilumab administration; Green diamonds refer to transfer towards medical wards and light blue diamond refers to discharge from the hospital. Black stars indicate repetition of infusion of Sarilumab during the follow-up. B) Rate of clinical response to Sarilumab in SARS-CoV-2^pos^ patients within 14 days follow-up (24h, 3,5,7 and 14 days) in ICU.

### Clinical course and safety profile in SARS-CoV-2^pos^ patients treated with Sarilumab

#### Medical-Wards

among 39 treated patients, 7(17.9%) required ICU admission, of whom 4(57.1%) were readmitted to the ward after 5-8 days and, at the time of manuscript submission, 1 patient was still hospitalized in ICU, whereas 2 patients died in ICU (one with chemical peritonitis due to duodenal perforation, not attributable to Sarilumab). (**Figure 1A**).

As shown in **Figure 1B**, the clinical improvement occurred in 18(46.2%) patients after 24h after Sarilumab infusion, while response rate at 3 days was 64.1%, reaching 89.7% at the last observation (median follow-up time of 16 days, range14-24). Twenty-four (70.6%) were discharged after a median of 12 days (range 5-22) after the first Sarilumab infusion. In addition, 30(85.7%) were no longer needing oxygen supplementation, 3(8.8%) were still at FiO_2_ 24%, 1(2.8%) at FiO_2_ 35% and 1(2.8%) in HFNC (FiO_2_ 35%), respectively. After Sarilumab infusion there was a progressive increase of PaO_2_/FiO_2_ ratio in SARS-CoV-2^pos^ patients (**Figure 1C**) mirroring the improvement of pulmonary inflammation at CT-scan (**Figure 1D-E**). A patient died for pulmonary embolization 5 days after Sarilumab administration.

#### ICU

Among 14 patients in ICU, 9(64.3%)(4 assisted with endotracheal intubation) were discharged to the medical wards within 5-12 days after Sarilumab administration (median 6.5 days). At the last observation, 8(88.9%) patients discharged to medical wards were no longer needing oxygen supplementation (4 of them discharged at home) and 1(11.1%) was still in HFNC (FiO_2_ 26%), while 5(35.7%) patients were still alive in ICU (4 significantly improved and one stable)(**Figure 2A-B**).

Overall mortality rate was 5.7% after Sarilumab administration: 1(2.5%) patient died in the Medical Ward whilst 2(14.2%) patients died in ICU, respectively.

**Supplementary Table 4** summarizes the adverse events in SARS-CoV-2^pos^ patients treated with Sarilumab.

### Predictors of response to Sarilumab treatment in SARS-CoV-2^pos^ patients

Among patients treated in medical wards, 25(64.1%) had clinical improvement 3 days after Sarilumab infusion. SARS-CoV-2^pos^ patients who improved were younger [63(53-73) years], with better respiratory parameters [PaO_2_/FiO_2_:175.9(133.4-229.2)] and with lower neutrophil/lymphocyte ratio [5.1(4.2-10.0)] than patients who remained unchanged or worsened at the same time point [age:74.0(67.5-77.3) years, p=0.01; PaO_2_/FiO_2_:127.3(99.5-147.6), p=0.004; N/L ratio: 8.2(6.1-11.9), p=0.03]. Moreover, patients who improved 3 days after Sarilumab administration had lower pre-treatment IL6 plasma levels [51.6(25.6-69.5) pg/ml] than patients who remained unchanged or worsened [120.4(88.2-158.4) pg/ml, p=0.001] (**Figure 3A-B, Supplementary Figure 1)**.

**Figure 3A-H.**
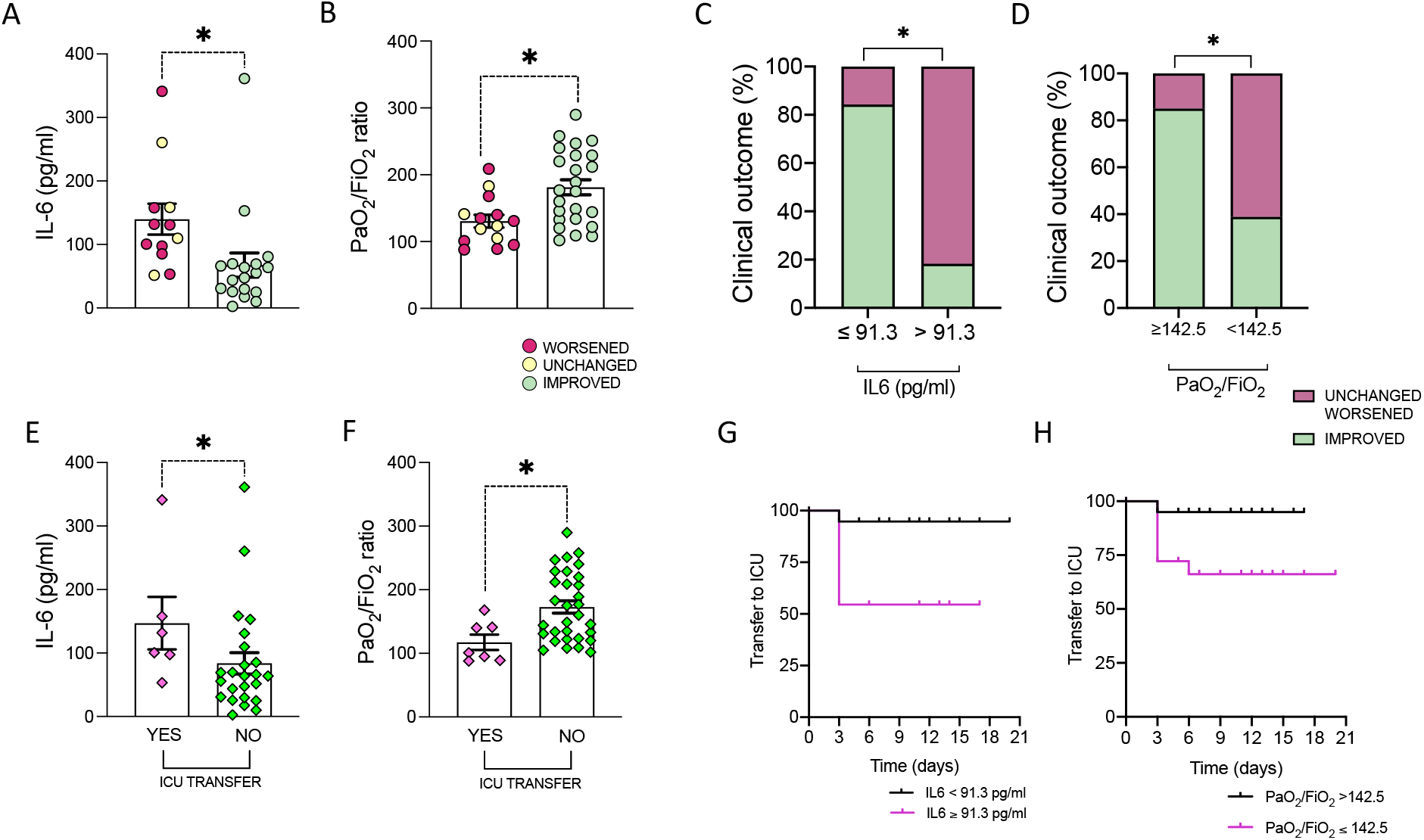
Clinical and biological baseline predictors of clinical outcome in SARS-Cov-2^pos^ patients treated with Sarilumab. **A)** Baseline IL-6 plasma levels in SARS-Cov-2^pos^ patients treated with Sarilumab based on clinical improvement at 3 days after Sarilumab treatment, *p=0.001. **B)** Baseline PaO_2_/FiO_2_ in SARS-Cov-2^pos^ patients treated with Sarilumab based on clinical improvement at 3 days after Sarilumab treatment, *p=0.004. **C)** Rate of clinical improvement at 3 days in SARS-Cov-2^pos^ patients treated with Sarilumab based on baseline IL-6 plasma levels, *p<0.0001. **D)** Rate of clinical improvement at 3 days in SARS-Cov-2^pos^ patients treated with Sarilumab based on baseline PaO_2_/FiO_2_, p=0.003. **E)** Baseline IL-6 plasma levels in SARS-Cov-2^pos^ patients based on transfer towards ICU within 14 days after after Sarilumab treatment, *p=0.05. **F)** Baseline PaO_2_/FiO_2_ in SARS-Cov-2^pos^ patients based on transfer towards ICU within 14 days after after Sarilumab treatment, *p=0.008. **G)** Kaplan-Meier analysis of transfer to ICU rate in SARS-Cov-2^pos^ patients treated with Sarilumab based on pre-treatment IL-6 plasma levels, p=0.01. **H)** Kaplan-Meier analysis of transfer to ICU rate in SARS-Cov-2^pos^ patients treated with Sarilumab based on pre-treatment PaO_2_/FiO_2_, p=0.03.

ROC curve analysis revealed that PaO_2_/FiO_2_ ratio≥142.5 and IL6 plasma levels≤91.3 pg/ml identifies SARS-CoV2^pos^ patients more likely to achieve clinical improvement at 3 days from Sarilumab administration (**Figure 3C-D, Supplementary Figure 2**).

**Table 1** shows the baseline characteristics of SARS-CoV-2^pos^ patients treated with Sarilumab in medical wards according to the need of ICU transfer, revealing that patients transferred to ICU had higher baseline IL-6 plasma levels (p=0.05) and lower PaO_2_/FiO_2_ ratio (p=0.007) than patients who did not need it (**Figure 3E-F**).

**Table 1.** Baseline demographic, clinical and inflammatory parameters of patients with SARS-Cov-2 pneumonia in medical ward setting, according to the need of ICU admission.

| Variables | Ward setting-treated patients<br>(n=39) |  | p |
| --- | --- | --- | --- |
|  | No ICU care<br>n=32 | ICU care<br>n=7 |  |
| Age, years [median(IQR)] | 64.5 (54.3-73.8) | 75.0 (71.0-79.0) | 0.01 |
| Male, n(%) | 30 (93.8) | 6 (85.7) | 0.47 |
| Weight, Kg [median(IQR)] | 80 (70.0-83.0) | 84 (75.0-100.0) | 0.21 |
| Overweight/obese, n(%) | 17 (53.1) | 6 (85.7) | 0.11 |
| **SIGNS AND SYMPTOMS** |  |  |  |
| Fever, n(%) | 39 (100.0) | 14 (100.0) | 1.00 |
| Cough, n(%) | 20 (51.3) | 9 (64.3) | 0.40 |
| Dyspnoea, n(%) | 23 (59.0) | 8 (57.1) | 0.91 |
| Days from symptoms to SARI infusion, [median(IQR)] | 11 (9.0-15.0) | 8 (3.0-11.0) | 0.06 |
| FiO<sub>2</sub> | 40 (35.0-50.0) | 60 (40.0-60.0) | 0.04 |
| PaO<sub>2</sub>/FiO<sub>2</sub> | 167.5 (125.4-226.5) | 101.0 (89.0-141.0) | 0.007 |
| **LABORATORY FINDINGS** |  |  |  |
| White blood cells count, x10<sup>3</sup>/ml [median(IQR)] | 6.1 (4.7-6.8) | 7.1 (5.7-10.1) | 0.22 |
| Lymphocytes count, x10<sup>3</sup>/ml [median(IQR)] | 0.8 (0.6-0.9) | 0.7 (0.5-0.9) | 0.38 |
| Neutrophils/Lymphocytes ratio, [median(IQR)] | 5.7 (4.2-11.3) | 8.0 (6.3-12.5) | 0.22 |
| Haemoglobin, g/L [median(IQR)] | 13.0 (11.8-13.9) | 13.3 (12.3-13.5) | 0.91 |
| Platelet, x10<sup>3</sup>/ml [median(IQR)] | 304 (247-420) | 271 (246.3-303.8) | 0.30 |
| Albumin, g/L [median(IQR)] | 2.9 (2.7-3.1) | 3.1 (2.7-3.8) | 0.28 |
| D-dimers, mg/L [median(IQR)] | 1345 (1056.5-2702) | 1955 (1108.3-3828.5) | 0.64 |
| GPT, mg/L [median(IQR)] | 32.5 (22.5-48.0) | 34.0 (17.0-48.0) | 0.82 |
| Creatinine, mg/L [median(IQR)] | 0.9 (0.7-1.2) | 1.2 (0.7-1.5) | 0.33 |
| C-Reactive Protein, mg/L [median(IQR)] | 148.5 (59.2-200.1) | 132.0 (102.8-161.5) | 0.78 |
| Lactate Dehydrogenase, [median(IQR)] | 315 (245.5-398.3) | 350 (306.5-469.0) | 0.24 |
| **SOLUBLE BIOMARKERS** |  |  |  |
| IL-6 plasma levels, pg/ml [median(IQR)] | 63.9 (30.0-103.7)*24 | 116.3 (86.3-203.7)*6 | 0.05 |
| **COMORBIDITY** |  |  |  |
| Diabetes, n(%) | 6 (18.8) | 1 (14.3) | 0.78 |
| Hypertension, n(%) | 15 (46.9) | 6 (85.7) | 0.06 |
| Cardiovascular disease, n(%) | 5/28 (17.9) | 2/6 (33.3) | 0.40 |
| Chronic Obstructive pulmonary disease, n(%) | 3/28 (10.7) | 1/6 (16.7) | 0.55 |
| Malignancy, n(%) | 0/28 (0.0) | 0/6 (0.0) | 1.00 |
| Dyslipidemia, n(%) | 6/29 (20.7) | 2/6 (33.3) | 0.50 |
| Chronic Infection, n(%) | 3/28 (10.7) | 0/6 (0.0) | 0.40 |

ICU: Intensive Care Unit; IQR: interquartile range; SARI: sarilumab; FiO<sub>2</sub>: fractional inspired oxygen; PaO<sub>2</sub>: arterial oxygen partial pressure; GPT: glutamine-pyruvate transaminase; IL: interleukin; g/L: grams/liter. \*number of patients with available IL-6 value.

ROC curve analysis revealed that PaO_2_/FiO_2_ ratio≤142.5 and IL6 plasma levels ≥91.3 pg/ml significantly identify SARS-CoV2^pos^ patients more likely to need to be transferred towards ICU within 14 days after Sarilumab treatment (**Figure 3G-H, Supplementary Figure 2**).

### Development of nomogram for the prediction of transfer to ICU in SARS-CoV-2^pos^ patients treated with Sarilumab in medical wards

Assessing the combined fulfillment of pre-treatment demographic (age), clinical (PaO_2_/FiO_2_) and laboratory parameters (IL6 plasma levels) in SARS-CoV2^pos^ patients treated with Sarilumab in medical wards, patients older than 75 years and having PaO_2_/FiO_2_≤142.5 had 40% incidence rate to be transferred to ICU. Moreover, patients with IL6 plasma levels≥91.3 pg/ml and PaO_2_/FiO_2_≤142.5 had 57.1% incidence to be transfer to ICU and that rate increases to 75% if older than 75 years (**Figure 4A-B**).

**Figure 4A-D.**
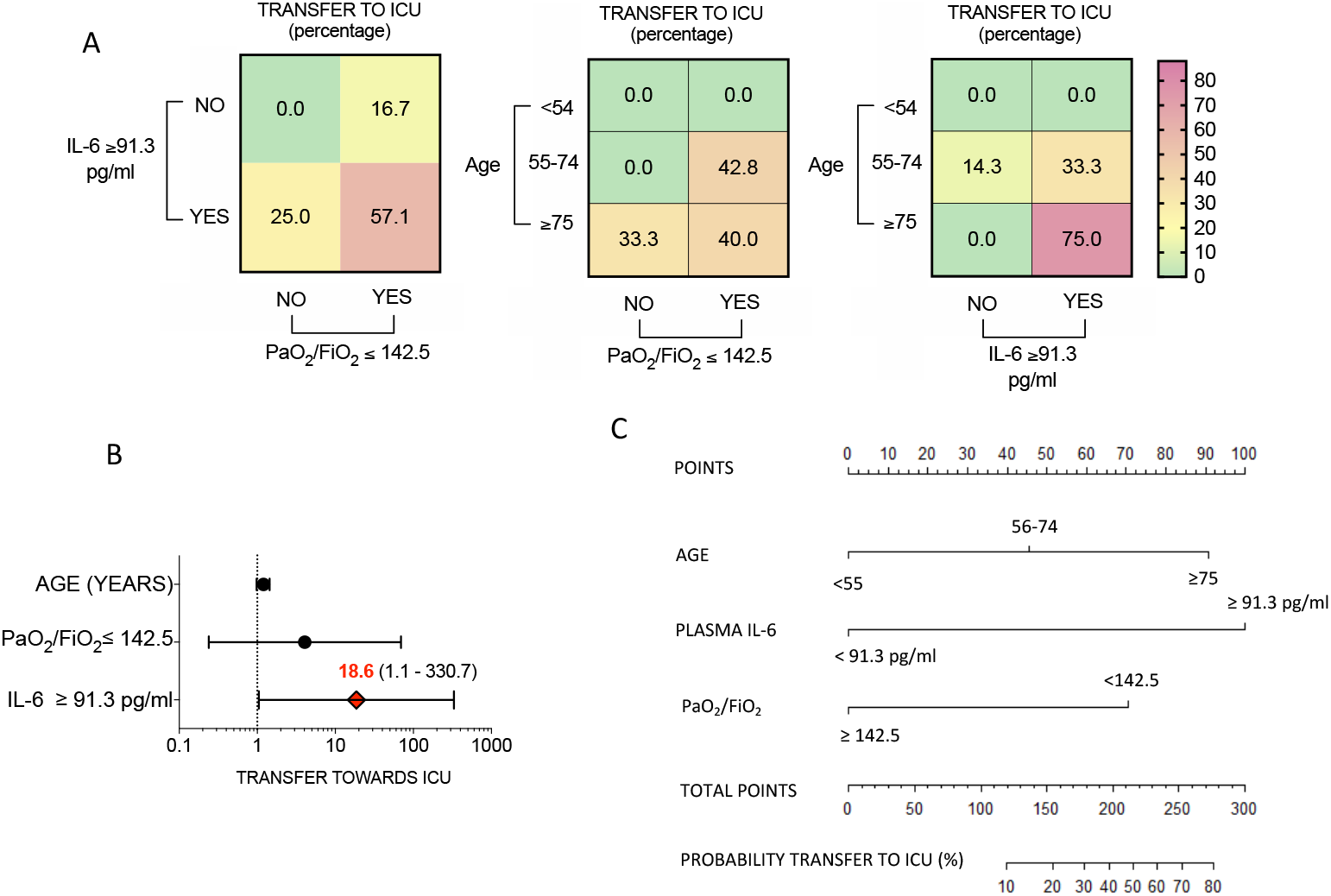
Development of multi-parametric nomogram predictive of transfer towards ICU in SARS-CoV-2^pos^ patients treated with Sarilumab in medical ward setting. **A)** Rate of transfer to ICU in SARS-CoV-2^pos^ patients treated with Sarilumab in medical wards based on pretreatment IL-6 plasma levels (91.3 pg/ml), PaO_2_/FiO_2_ ratio value (142.5) and age category (<54 years, 55-74 years and >75 years respectively); Values are expressed as percentages. **B)** Odd ratios of transfer to ICU in SARS-CoV-2^pos^ patients treated with Sarilumab in medical wards based on the fulfillment of pre-treatment definite parameters; Values are expressed as Odd Ratio (95%CI); **C)** Nomogram for the computation of probability of transfer to ICU in SARS-CoV-2^pos^ patients treated with Sarilumab in medical wards. **IL**: Interleukin; **ICU**: Intensive Care Unit.

A nomogram was created (**Figure 4C**) incorporating the three significant risk factors predicting the “transfer to ICU”. The value of each variable was given a score on the “points” scale axis. A total score was calculated by adding each single point score and by projecting the value of the “total points” score to the lower “Probability” line.

## DISCUSSION

Reports of different cohorts stated that nearly 80% of SARS-CoV-2 infected subjects have a spontaneous recovery, while 15% experience symptoms needing hospitalization and up to 5% may require care within ICU. The most aggressive phase is thought to have an aggressive inflammatory background (cytokine release syndrome-CRS) that leads to the respiratory distress.^4^ At this stage, IL-6 is a key player in regulating such cascade^10^ and SARS-CoV-2^pos^ patients show increased circulating IL-6 levels associated with subsequent development of cytokines release syndrome.^13^ Therefore, IL-6R inhibitors (i.e. Tocilizumab or Sarilumab) may help to prevent disease progression and promote faster resolution of SARS-CoV-2 pneumonia.^10,13-15^ Thus, in China, nearly 200 patients with severe SARS-CoV-2 pneumonia were treated with Tocilizumab (source: Roche Ltd, China) and, although the results of this study have not been made public to date, the Chinese CDC, approved Tocilizumab use in patients with SARS-CoV-2 severe pneumonia needing to be admitted to ICU.^16^ These indications were also implemented in the SIMIT Lombardy guidelines released on March 20^th^, 2020. Given the emerging evidence on the efficacy of Tocilizumab in patients with severe pneumonia, currently this drug is used even in a medium-severe phase.

The ensuing shortage of Tocilizumab led COVID-19 Task Force of our Hospital to adopt the option of using Sarilumab as an alternative anti-IL-6R. At the time of manuscript submission, there were 8 registered controlled trials testing Sarilumab as possible effective alternative to treat SARS-CoV-2 severe pneumonia.^17^

Herein, we report, for the first time, the clinical outcome and safety profile of Sarilumab in SARS-CoV-2 severe pneumonia both in medical wards and ICU setting. Sarilumab is available in subcutaneous formulation which is administered every 2 weeks in patients with RA^18^ showing 20 times higher affinity for IL6R (both soluble and membrane-bound IL6 receptors) than Tocilizumab.^19^

Our data show that Sarilumab has a good global rate of clinical efficacy to treat SARS-CoV-2 severe pneumonia in terms of clinical improvement and mortality. Overall, 83.0% of 53 SARS-CoV2^pos^ patients (89.7% in medical wards and 64.3% in ICU) safely resolved their severe respiratory syndrome, with an overall mortality rate of 5.7%. Moreover, even considering patients with a critical lung involvement, Sarilumab treatment led up to 92.8% clinical improvement with an ICU discharge rate of 64.3% in a mean recovery time of 7 days. Interestingly, only 2 patients were treated with a short course of GC. Avoiding secondary infections due to GC is clinically relevant, also in light of some preliminary negative results of corticosteroid use in treatment of Covid-19 patients with lung injury.^20^

Moreover, Sarilumab appears to be safe and we did not register drug-related serious adverse events and no secondary infections up to the last follow-up.

Initial reports revealed that baseline demographic and clinical parameters may be useful to assess SARS-CoV-2 infection prognosis.^21^ In this contest, we found that young SARS-CoV-2^pos^ patients with severe pneumonia with PaO_2_/FiO_2_ ratio>142.7 and not extreme systemic inflammation (IL-6 plasma levels<91.3 pg/ml) have the highest chance of Sarilumab efficacy in terms of early clinical response and less need to be transferred to ICU than patients not fulfilling these criteria. Whether a higher intensity drug exposure (more i.v. infusions) would better control the more aggressive underlying inflammation, will be highligthed only by a targeted randomized trial using a biomarker-oriented approach. Conversely, a multiparametric nomogram including such variables allows to predict the need of being transferred to ICU up to 80% of patients with severe pneumonia. Interestingly, analysing the dynamics of clinical response of patients treated with Sarilumab, it arose that the sooner therapy is started, the sooner the positive result in terms of clinical improvement is observed (“window of opportunity”). In this study neither D-Dimers levels, nor lymphopenia arose as predictors of a critical outcome, while the pre-treatment PaO/FiO_2_ value had the greatest role along with age.

The limitations of this report are its open-label nature with no comparison group, the limited number of critical-ICU patients and adoption of concomitant treatments of unproven efficacy (possibly increasing the safety results). Moreover, a longer follow-up may be useful to increase the strength of our findings in terms of long-term efficacy and safety of Sarilumab in SARS-CoV-2 severe pneumonia. Yet, an outpatient clinic for these patients has already been set-up to collect prospective clinical and biological informations.

In conclusion, IL-6R inhibition is a successful treatment strategy for severe SARS-CoV-2 pneumonia and Sarilumab is a valid and safe alternative in the therapeutic armamentarium of this disease without defined standardized treatment algorythms.

## Data Availability

N.A.

## Aknowledgements

We would like to thank all the patients who sadly were infected by SARS-CoV-2 during this pandemic and participated to the study. We would like to thank Prof. Rocco Bellantone, Dr. Andrea Cambieri, Prof. Marco Elefanti and Prof. Giovanni Scambia of the Fondazione Policlinico Universitario A. Gemelli IRCCS for continuous support to all the activities of the FPG-IRCCS COVID-19 multidisciplinary Task-Force.

## Conflict of interest statement

All authors declare no conflict of interest with the submitted manuscript.

**Supplementary Table 1.** Demographic and clinical characteristics of 39 patients with severe SARS-CoV-2 pneumonia treated with Sarilumab in medical wards.

| Patient ID | Sex | Age | Obesity | N. of comorbidities | PaO<sub>2</sub>/FiO<sub>2</sub> | IL6 (pg/ml) | Time from symptoms to SARI treatment (days) |
| --- | --- | --- | --- | --- | --- | --- | --- |
| #1 | M | 71 | Overweight | Dysl, COPD | 101 | 97,42 | 8 |
| #2 | M | 74 | Overweight | AH, IC | 133 | 30,90 | 15 |
| #3 | M | 51 | Normal weight | AH, IC | 109 | 55,66 | 11 |
| #4 | M | 75 | Normal weight | - | 105 | 260,53 | 7 |
| #5 | M | 63 | Obese | AH, DM | 146 | 63,90 | 9 |
| #6 | M | 73 | Normal weight | AH, Dysl, IC | 140 | 53,11 | 13 |
| #7 | M | 40 | Normal weight | - | 144 | 69,44 | 11 |
| #8 | M | 63 | Overweight | AH, Dysl, IC | 251 | 66,25 | 10 |
| #9 | M | 66 | Normal weight | AH, DM, Dysl, IC, CI | 247 | n.a. | 5 |
| #10 | M | 77 | Overweight | AH | 168 | 157,90 | 6 |
| #11 | M | 66 | Overweight | AH | 119 | 109,89 | 16 |
| #12 | M | 78 | Overweight | - | 131 | 85,17 | 4 |
| #13 | M | 73 | Overweight | AH | 108 | 43,88 | 5 |
| #14 | M | 55 | Normal weight | AH, Dysl | 212 | 17,58 | 11 |
| #15 | M | 73 | Obese | - | 122 | n.a. | 5 |
| #16 | M | 60 | Normal weight | AH, IC | 209 | 130,88 | 11 |
| #17 | M | 80 | Overweight | AH, COPD | 88 | 341,23 | 10 |
| #18 | F | 50 | Normal weight | - | 189 | n.a. | 11 |
| #19 | F | 79 | Overweight | AH | 95 | 131,98 | 11 |
| #20 | M | 68 | Overweight | Dysl. | 183 | 158,53 | na |
| #21 | M | 54 | Normal weight | - | 290 | 69,57 | 11 |
| #22 | M | 75 | Obese | AH, DM | 141° | n.a. | 3 |
| #23 | M | 68 | Overweight | AH, DM, Dysl, COPD | 207 | 24,91 | 18 |
| #24 | M | 69 | Obese | - | 248* | 47,55 | 5 |
| #25 | M | 48 | Overweight | - | 229 | 29,70 | 18 |
| #26 | M | 76 | Normal weight | AH | 175 | 63,99 | 20 |
| #27 | M | 75 | Normal weight | AH, CI | 123 | 51,45 | 13 |
| #28 | M | 71 | Overweight | AH, DM | 102* | 153,01 | 12 |
| #29 | M | 55 | Normal weight | - | 229 | 10,04 | 9 |
| #30 | M | 50 | Normal weight | AH | 160 | n.a. | 9 |
| #31 | M | 54 | Overweight | - | 258 | 361,39 | 16 |
| #32 | M | 87 | Overweight | AH, COPD | 177 | 80,71 | 12 |
| #33 | F | 52 | Obese | - | 220 | 25,78 | 11 |
| #34 | M | 59 | Normal weight | DM | 240 | 2,52 | 14 |
| #35 | M | 70 | Obese | AH, IC | 89° | 100,68 | 1 |
| #36 | M | 55 | Overweight | - | 135 | n.a. | 8 |
| #37 | M | 55 | Normal weight | CI | 120 | n.a. | 10 |
| #38 | M | 77 | Overweight | DM, Dysl | 149 | n.a. | 19 |
| #39 | M | 95 | Normal weight | - | 134 | n.a. | 15 |

AH: Arterial Hypertension; DM: Diabetes; Dysl: dyslipidemia; IC: Ischemic Cardiopathy; CI: Chronic infections; na: not available. pg/ml: picograms/milliliter. PaO<sub>2</sub>/FiO<sub>2</sub>: arterial oxygen partial pressure/fraction of inspired oxygen. \*Continuous Positive Airway Pressure-CPAP; °Non-invasive ventilation-NIV.

**Supplementary Table 2.** Demographic and clinical characteristics of 14 patients with critical SARS-CoV-2 pneumonia treated with Sarilumab in ICU setting.

| Patient ID | Sex | Age | Obesity | N. of comorbidities | PaO<sub>2</sub>/FiO<sub>2</sub> | IL6 (pg/ml) | Time from symptoms to SARI treatment (days) |
| --- | --- | --- | --- | --- | --- | --- | --- |
| #1 | M | 72 | Normal weight | - | 110* | n.a. | 13 |
| #2 | M | 52 | Obese | - | 101* | 116,36 | 12 |
| #3 | M | 46 | Obese | DM | 170^ | 68,76 | 4 |
| #4 | M | 60 | Overweight | AH | 92* | 99,07 | 8 |
| #5 | M | 57 | Overweight | - | 120^ | 66,18 | 10 |
| #6 | M | 53 | Normal weight | - | 113^ | n.a. | 17 |
| #7 | M | 54 | Overweight | - | 105* | 66,75 | 13 |
| #8 | F | 68 | Overweight | AH,DM | 232* | n.a. | 5 |
| #9 | M | 68 | Overweight | AH,DM,Dysl, IC, neoplasm | 111* | n.a. | 19 |
| #10 | M | 61 | Obese | AH | 129^ | 120,33 | 12 |
| #11 | F | 65 | Normal weight | - | 79* | 47,85 | 11 |
| #12 | M | 69 | Overweight | AH,DM,Dysl. IC | 154* | 63,11 | 18 |
| #13 | F | 59 | Normal weight | Neoplasm | 99^ | 30,50 | 9 |
| #14 | M | 62 | Obese | AH,Dysl., IC | 297* | 89,44 | 13 |

AH: Arterial Hypertension; DM: Diabetes; Dysl: dyslipidemia; IC: Ischemic Cardiopathy; CI: Chronic infections; na: not available. pg/ml: picogram/milliliter. PaO<sub>2</sub>/FiO<sub>2</sub>: arterial oxygen partial pressure/fraction of inspired oxygen. \*ETI: Endo-tracheal Intubation; ^NIV: Non-invasive ventilation.

**Supplementary Table 3.** Demographic, clinical and inflammatory parameters of patients with SARS-Cov-2 infection enrolled in the study.

| Variables | Whole cohort<br>(n=53) |  |
| --- | --- | --- |
|  | Medical wards care<br>n=39 | ICU care<br>n=14 |
| Age, years [median(IQR)] | 68.0 (55.0-75.0) | 60.5 (53.8-68.0) |
| Male, n(%) | 36 (92.3) | 12 (85.7) |
| Weight, Kg [median(IQR)] | 80.0 (70.0-85.0) | 85.0 (70.0-90.0) |
| Overweight/obese, n(%) | 23 (59.0) | 10 (71.4) |
| **SIGNS AND SYMPTOMS** |  |  |
| Fever, n(%) | 39 (100.0) | 14 (100.0) |
| Cough, n(%) | 20 (51.3) | 9 (64.3) |
| Dyspnoea, n(%) | 23 (59.0) | 8 (57.1) |
| Days from symptoms to SARI infusion, [median(IQR)] | 11.0 (7.8-13.3) | 12.0 (8.8-14.0) |
| FiO<sub>2</sub> [median(IQR)] | 50.0 (35.0-60.0) | 50.0 (45.0 – 60.0) |
| PaO<sub>2</sub>/FiO<sub>2</sub> [median(IQR)] | 146.4 (120.0-212.0) | 113.3 (100.1-162.0) |
| **LABORATORY FINDINGS** |  |  |
| White blood cells count, x10<sup>3</sup>/ml [median(IQR)] | 6.2 (4.7-7.7) | 7.3 (5.9-10.2) |
| Lymphocytes count, x10<sup>3</sup>/ml [median(IQR)] | 0.8 (0.6-0.9) | 0.9 (0.5-1.4) |
| Neutrophils/Lymphocytes ratio, [median(IQR)] | 6.8 (4.6-11.7) | 8.2 (4.7-11.6) |
| Haemoglobin, g/L [median(IQR)] | 13.0 (12.1-13.8) | 12.3 (11.7-13.6) |
| Platelet, x10<sup>3</sup>/ml [median(IQR)] | 293 (252.0 – 404.0) | 288 (231.5-379.5) |
| Albumin, g/L [median(IQR)] | 2.9 (2.7-3.1) | 2.6 (2.4 -2.8) |
| D-dimers, mg/L [median(IQR)] | 1403.0 (1056.5 – 2702.0) | 1372.0 (1042.5-11321.3) |
| GPT, mg/L [median(IQR)] | 34.0 (19.5-47.5) | 52.0 (32.3-80) |
| Creatinine, mg/L [median(IQR)] | 0.9 (0.7-1.2) | 0.8 (0.7-1.0) |
| C-Reactive Protein, mg/L [median(IQR)] | 142.7 (68.3-186.1) | 169.7 (152.5-220.3) |
| Lactate Dehydrogenase, [median(IQR)] | 327.0 (269.5 - 410.8) | 426.0 (339.0-548.5) |
| **SOLUBLE BIOMARKERS** |  |  |
| IL-6 plasma levels, pg/ml [median(IQR)] | 67.9 (40.6 - 131.2)* | 67.8 (59.3-103.4)** |
| **ANY COMORBIDITY** |  |  |
| Diabetes, n(%) | 7 (17.9) | 4(28.6) |
| Hypertension, n(%) | 21 (53.8) | 6 (42.9) |
| Cardiovascular disease, n(%) | 7/34 (20.6) | 3/12 (25.0) |
| Chronic Obstructive pulmonary disease, n(%) | 4/34 (11.7) | 0/12 (0.0) |
| Malignancy, n(%) | 0/34 (0.0) | 2/12 (16.7) |
| Dyslipidemia, n(%) | 8/35 (22.9) | 3/12 (25) |
| Chronic Infection, n(%) | 3/34 (8.8) | 0/12 (0.0) |

ICU: Intensive Care Unit; IQR: interquartile range; SARI: sarilumab; GPT: glutamine-pyruvate transaminase; IL: interleukin. g/L: grams/liter; pg/ml: picograms/liter; FiO<sub>2</sub>: fractional inspired oxygen; PaO<sub>2</sub>: arterial oxygen partial pressure. \*30 patients with available IL-6 value; \*\*10 patients with available IL-6 value.

**Supplementary Table 4.** Adverse events recorded in patients with severe SARS-CoV-2 pneumonia treated with Sarilumab.

|  | **Frequency** | **Action** | **Solved N/Y** |
| --- | --- | --- | --- |
| **Adverse events** | 22(41.5%) | - | Y |
| **Laboratory** |  |  |  |
| **Mild neutropenia (>500/ul)** | 5(9.4) | None | Y |
| **severe neutropenia (< 500/ul)** | 3(5.7) | injection of GMCSF | Y |
| **Increased Liver enzymers (4 UNL)** | 6(18.8) | None | Y |
| **Mild thrombocytopenia** | 1 | None | Y |
| **Pulmonary Embolism** | 1 | Specific treatment | Y |
| **Deep Venous Thrombosis** | 2 | Specific treatment | Y |
| **Infections** | 0 | - | - |

**GMCSF:** Granulocyte-Macrophage Colony-Stimulating Factor; **UNL:** Upper Normal Limit; **N:** No; **Y:** Yes.

## Supplementary information about the author-list of the manuscript

### The authors of the present manuscript are on behalf of GEMELLI AGAINST COVID-19 Group

#### Gastroenterology and Internal Medicine

Abbate Valeria, Addolorato Giovanni, Agostini Fabiana, Ainora Maria Elena, Andriollo Gloria, Annicchiarico Brigida Eleonora, Antonelli Mariangela, Antonucci Gabriele, Armuzzi Alessandro, Bibbò Stefano, Bove Vincenzo, Buonomo Alessandro, Cammà Giulia, Caruso Cristiano, Casciaro Francesco Antonio, Cecchini Andrea Leonardo, Cerrito Lucia, Coppola Gaetano, D’Addio Stefano, D’Alessandro Alessia, D’Aversa Francesca, De Siena Martina, Del Zompo Fabio, Di Luca Roberta, Feliciani Daniela, Funaro Barbara, Gaetani Eleonora, Garcovich Matteo, Gasbarrini Antonio, Guidone Caterina, Iaconelli Amerigo, Ianiro Gianluca, Iaquinta Angela, Impagnatiello Michele, Landi Rosario, Leo Massimo, Liguori Antonio, Lopetuso Loris, Macerola Noemi, Mancarella Francesco, Mangiola Francesca, Marrone Giuseppe, Matteo Maria Valeria, Miele Luca, Mignini Irene, Milani Alessandro, Monti Flavia, Mora Vincenzina, Murace Celeste Ambra, Napoli Marco, Nardella Elisabetta, Nicoletti Alberto, Papa Alfredo, Papparella Luigi Giovanni, Paratore Mattia, Parrinello Giuseppe, Pero Erika, Pizzoferrato Marco, Pizzolante Fabrizio, Pompili Maurizio, Pontecorvi Valerio, Ponziani Francesca, Popolla Valentina, Porceddu Enrica, Pulcini Gabriele, Rapaccini Gian Ludovico, Rinninella Emanuele, Rossini Enrica, Rovedi Fabiana, Rumi Gabriele, Salvatore Lucia, Santini Paolo, Santopaolo Francesco, Sarnari Caterina, Schepis Tommaso, Schiavello Francesca, Sestito Luisa, Settanni Carlo, Stella Leonardo, Talerico Rossella, Tarli Claudia, Tosoni Alberto, Vetrone Lorenzo, Zileri Dal Verme Lorenzo

#### General Medicine

Amato Elena, Burzo Livia, Cianci Rossella, Costante Federico, De Matteis Giuseppe, De Vito Francesco, Di Gialleonardo Luca, Gambassi Giovanni, Porfidia Angelo, Rossi Raimondo

#### Endocrinology

Bianchi Antonio, Corsello Andrea, Del Gatto Valeria, Gelli Silvia, Giampietro Antonella, Locantore Pietro, Milardi Domenico, Policola Caterina, Rossi Laura, Zelano Lorenzo

#### Rheumatology Team

Alivernini Stefano, Bosello Silvia Laura, Bruno Dario, Fedele Anna Laura, Gigante Laura, Gremese Elisa, Lizzio Marco Maria, Natalello Gerlando, Paglionico Annamaria, Perniola Simone, Petricca Luca, Tolusso Barbara, Varriano Valentina, Verardi Lucrezia

#### Geriatrics

Acampora Nicola, Barillaro Christian, Bellieni Andrea, Benvenuto Francesca, Bernabei Roberto, Bramato Giulia, Brandi Vincenzo, Carfi Angelo, Ciciarello Francesca, Cipriani Maria Camilla, D’Angelo Emanuela, Falsiroli Cinzia, Fusco Domenico, Landi Francesco, Landi Giovanni, Liperoti Rosa, Lo Monaco Maria Rita, Martis Ilaria, Martone Anna Maria, Marzetti Emanuele, Pagano Francesco Cosimo, Pais Cristina, Rocchi Sara, Rota Elisabetta, Russo Andrea, Salerno Andrea, Salini Sara, Tosato Matteo, Tritto Marcello, Tummolo Anita Maria, Zuccalà Giuseppe

#### Medical Clinic

Biasucci Luigi Marzio, Biscetti Federico, Capristo Esmeralda, Flex Andrea, Gallo Antonella, Landolfi Raffaele, Montalto Massimo, Nesci Antonio, Pecorini Giovanni, Pola Roberto, Santoliquido Angelo

#### Nuerologist

Guglielmi Valeria, Iorio Raffaele, Nicoletti Tommaso, Tricoli Luca

#### Emercency Medicine

Akacha Karim, Barone Fabiana, Benicchi Andrea, Bonadia Nicola, Bosco Giulia, Bungaro Maria Chiara, Candelli Marcello, Capaldi Lorenzo, Carbone Luigi, Cardone Silvia, Carnicelli Annamaria, Cicchinelli Sara, Covino Marcello, De Cunzo Tommaso, De Luca Giulio, Della Polla Davide, Di Maurizio Luca, Esperide Alessandra, Forte Evelina, Franceschi Francesco, Franza Laura, Fuorlo Mariella, Gabrielli Maurizio, Gasparrini Irene, Giuliano Giorgia, Giupponi Bianca, Kadhim Cristina, Loria Valentina, Manno Alberto, Marchesini Debora, Migneco Alessio, Navarra Simone Maria, Nicolò Rebecca, Nista Enrico Celestino, Nuzzo Eugenia, Ojetti Veronica, Petrucci Martina, Piccioni Andrea, Pignataro Giulia, Privitera Giuseppe, Racco Simona, Romano Stefano, Rosa Federico, Sabia Luca, Samori Dehara, Santarelli Luca, Santoro Michele Cosimo, Sardeo Francesco, Saviano Angela, Saviano Luisa, Simeoni Benedetta, Tilli Pietro, Torelli Enrico, Valletta Federico, Zaccaria Raffaella

#### Pulmonology Unit

Baldi Fabiana, Berardini Ludovica, Bruni Teresa, Calabrese Angelo, Calvello Maria Rosaria, Corbo Giuseppe Maria, Fiore Maria Chiara, Inchingolo Riccardo, Intini Erica, Giuliano, Pasciuto Giuliana, Porro Lucia Maria, Potenza Annalisa, Richeldi Luca, Siciliano Matteo, Simonetti Jacopo, Smargiassi Andrea, Varone Francesco

#### Infectious Diseases

Borghetti Alberto, Cauda Roberto, Ciccullo Arturo, Cingolani Antonella, Damiano Fernando, De Gaetano Kathleen, Del Giacomo Paola, Di Giambenedetto Simona, Dusina Alex, Faliero Domenico, Fantoni Massimo, Giuliano Gabriele, Izzi Immacolata Maria, Losito Angela Raffaella, Lucia Mothanje Barbara Patricia, Maiuro Giuseppe, Moschese Davide, Siciliano Valentina, Murri Rita, Pallavicini Federico, Picarelli Chiara, Raffaelli Francesca, Scoppettuolo Giancarlo, Siciliano Valentina, Taddei Eleonora, Tamburrini Enrica, Tumbarello Mario, Ventura Giulio, Visconti Elena

#### Intensive Care Unit

Anzellotti Gianmarco, Andreani Francesca, Annetta Maria Giuseppina, Barattucci Ilaria, Bello Giuseppe, Biasucci Daniele Guerino, Bini Alessandra, Bisanti Alessandra, Bocci Maria Grazia, Bongiovanni Filippo, Cambise, Chiara, Canistro Gennaro, Cantanale Antonello, Capone Emanuele, Carelli Simone, Cesarano Melania, Chiarito Annalisa, Cicetti Marta, Consalvo Maria Ludovica, Costanzi Matteo, Crupi Davide, Cutuli Salvatore Lucio, De Berardinis Gianmaria, De Pascale Gennaro, De Santis Paolo, Dell’Anna Antonio Maria, Di Muro Mariangela, Eleuteri Davide, Fachechi Daniele, Ferrante Cristina, Ferrone Giuliano, Festa Rossano, Francesconi Alessandra, Galletta Claudia, Grieco Domenico Luca, Guerrera Manuel, Gullì Antonio, Iafrati Aurora, Jovanovic Tamara, La Macchia Rosa, Levantesi Laura, Lombardi Gianmarco, Maggi Giampaolo, Maresca Gianmarco, Mattana Chiara, Maviglia Riccardo, Memoli Carmen, Montini Luca, Morena Tony Christian, Murdolo Martina, Natalini Daniele, Nicoletti Rocco, Oggiano Marco, Paiano Gianfranco, Paolillo Federico, Papanice Domenico, Pasculli Pierpaolo, Piccolo Annalisa, Piervincenzi Edoardo, Pintaudi Gabriele, Pisapia Luca, Pontecorvi Flavia, Pozzana Francesca, Romanò Bruno, Rubino Carlotta, Russo Andrea, Sandroni Claudio, Santolamazza Danilo, Scarascia Roberta, Sedda Davide, Sessa Flaminio, Sicuranza Rossella, Soave Paolo Maurizio, Sonnino Chiara, Stella Claudia, Stival Eleonora, Tamburello Elio, Tanzarella Eloisa Sofia, Tarascio Elena, Tersali Alessandra, Timpano Jacopo, Torrini Flavia, Vallecoccia Maria Sole, Vassalli Francesco, Viviani Andrea, Zaccone Carmelina

#### Crisis Unit

Bellantone Rocco Domenico Alfonso, Berloco Filippo, Cambieri Andrea, Capalbo Gennaro, Catalano Lucio, D’alfonso Maria Elena, La Milia Daniele Ignazio, Nicolotti Nicola, Pignataro Raffaele, Sanguinetti Maurizio, Staiti Domenico, Vetrugno Giuseppe

